# High Initial Titres of Anti-Spike Antibodies following SARS-CoV-2 Infection is Associated with Faster Decay Rates at Four Months Follow-Up

**DOI:** 10.1101/2021.03.02.21252362

**Authors:** Vidya Menon, Masood A Shariff, Victor Perez Gutierrez, Juan M Carreño, Bo Yu, Muzamil Jawed, Marcia Gossai, Elisenda Valdez, Anjana Pillai, Usha Venugopal, Moiz Kasubhai, Vihren Dimitrov, Florian Krammer

## Abstract

**Background:** Dynamics of humoral immune responses to SARS-CoV-2 antigens following infection suggests an initial decay of antibody followed by subsequent stabilization. We aim to understand the longitudinal humoral responses to SARS-CoV-2 nucleocapsid (N) protein and spike (S) protein and to evaluate their correlation to clinical symptoms among healthcare workers (HCW).

**Methods:** In this cross-sectional longitudinal cohort study done in two phases over four months, HCW underwent serial qualitative serology testing for anti-N antibody, quantitative MSH-ELISA to detect Receptor Binding Domain and full-length S reactive antibodies and completed online surveys about COVID-19 related symptoms and healthcare/community exposure.

**Results:** Anti-N antibody positivity was 27% and anti-S positivity was 28% in Phase 1. In Phase 2 anti-S titres were higher in symptomatic than in asymptomatic positive subjects in Phase 1. Marginally higher titers were seen in asymptomatic compared to the symptomatic positive subgroup in Phase 2. A positive correlation was noted between age, number and duration of symptoms, and Phase 1 anti-S antibody titre. A strong correlation was observed between Phase 1 titers and decay of anti-S antibody titres between the two phases. Significant correlation with rate of decay was also noted with fever, GI symptoms, and total number and duration of COVID-19 symptoms.

**Conclusions:** Higher initial anti-S antibody titres were associated with larger number and longer duration of symptoms as well as faster decay during the two time points.

**Key Points:** *Question:* What is the decay rate of neutralizing antibodies among SARS-CoV-2 infected healthcare workers?

*Findings:* In this cohort study that included 178 healthcare workers, over a 4-month period following the COVID-19 pandemic, participants had an initial rise in anti-nucleocapsid (N) and anti-spike (S) antibodies, which was followed by decay and stabilization of the titres. Significant correlation with rate of decay was noted with the symptomatic participants.

*Meaning:* A strong correlation is observed in the decay of anti-S antibody titres based on symptomology, thus eluding to the fact that continued recommendations for infection protection and COVID-19 vaccine campaigns are necessary.

## Introduction

In light of the unprecedented coronavirus disease 2019 (COVID-19) pandemic, understanding the role of the immune system in countering the viral infection is critical not just to design effective antiviral strategies but also to aid us in taking appropriate public health decisions. The early publication of the viral genome led to a rapid development of many nucleic acid based diagnostic assays for severe acute respiratory syndrome coronavirus 2 (SARS-CoV-2) infections. While nucleic acid-based tests are widely employed in the diagnosis of acute (current) SARS-CoV-2 infections, they are often limited in their clinical utility in identifying past infections or assess the level of immunity to SARS-CoV-2 within the communities. Evaluation of antibody responses is the other well-known modality used in a clinical setting that can detect both current, and past infections and is the preferred approach for surveillance to determine the true prevalence of infections. The currently available serological assays for SARS-CoV-2 target either the viral nucleoprotein (N) or the spike surface protein (S) antigens. The S-protein, which contains the receptor binding domain (RBD), binds to host cells via the angiotensin converting-enzyme-2 (ACE2) receptor, followed by membrane fusion^1,2^. The spike is the target of most neutralizing antibodies ^3–5^, while the N plays an important role in transcription enhancement and viral assembly ^6^. Studies have demonstrated that antibodies against the N and S appeared around the same time - between day 8 and day 14 after the onset of symptoms with antibodies to the N being more sensitive than anti S antibodies for detecting early infection^7^. Neutralizing antibodies confer protective immunity and can be detected in most infected individuals 10-15 days following the onset of COVID-19 symptoms and remain elevated following initial viral clearance ^8–12^. There is compelling evidence suggesting that serological assays for anti-S antibodies predict neutralizing activity, in contrast to N based assays^11,13^.

The detailed characterization of the dynamics of humoral immune responses to the SARS-CoV-2 viral antigens following infection is still ongoing and early evidences suggest an initial decay of antibody followed by stabilization at a certain level^11,14–18^. These dynamics are likely driven by an initial expansion of plasmablasts which produce large amounts of antibody but die off quickly followed by a slower decay of antibody titres (the half-life of IgG is approximately three weeks) which then transitions into a steady state level of antibody produced by long-lived plasma cells^19^. However, it is currently unknown, if the magnitude of the initial expansion of plasmablast and the associated antibody titres are correlated with the steady state level of serum antibody produced by long-lived plasma cells. This is an important question since steady state antibody levels may provide superior protection from re-infection^20,21^.

Specifically, there is currently a paucity of information on the kinetics of antibody decay among health care workers (HCW). It is suspected that SARS-CoV-2 infections among HCW are usually asymptomatic or mildly symptomatic and frequently associated with either underreporting of symptoms or heterogenous PCR and/or serologic diagnostics leading to most of them going undetected or unrecognized^22^. A large cohort study of HCWs in the greater New York City (NYC) area showed a seroprevalence of SARS-CoV-2 antibodies of 13.7%^23^. Our own data of anti N antibody screening among HCW at a New York City public hospital in the Bronx following the first “surge” of COVID-19 in May 2020, found that SARS-CoV-2 seroprevalence was at 27%^24^. Understanding the longitudinal kinetics of SARS-CoV-2 antibody response and the effectiveness of commercial antibody measurement assays is crucial to correctly determine infection rates, sero-prevalence and true sero-reversion rates in both infected and vaccinated individuals – and to better understand protection associated with sero-positivity.

In this study, we aimed to investigate the longitudinal humoral responses to viral N and the spike and to evaluate their correlation to clinical symptoms and baseline characteristics in our HCW study cohort. Importantly, having access to samples during the initial antibody peak and several months out, we also aimed to determine if initial high antibody levels correlated with high antibody titers at steady state.

## Methods

### Study setting and population

The study was a cross sectional cohort study done in two phases after receiving Institutional Review Board approval (IRB # 20-009, Lincoln Medical Center, Office of the Institutional Review Board approved as per 45 CFR 46 & 21 CFR50,56 under a full board committee and gave its approval on 4/28/2020). The Phase 1 was conducted in May/June, 2020 and the Phase 2 was completed August/September 2020. The cohort included HCWs who worked at the New York City Public Hospital in the South Bronx and were willing to participate in both phases of the study. In the Phase 1 of the study, after informed consent, participants underwent qualitative serology testing (Abbott Architect SARS-CoV-2 IgG Assay, Abbott Park, IL 60064 USA)^25^ and a nasopharyngeal swab for SARS-CoV-2 (Bio-Reference Laboratories, Inc., Elmwood Park, NJ, USA). They also completed an initial online survey on demographics, symptoms of COVID-19, healthcare/community exposure etc. An extra sample was collected and stored at −80°C for subsequent analysis. These samples were processed using a quantitative enzyme-linked immunosorbent assay (ELISA) that correlates well with virus neutralization, developed by Mount Sinai Health System (MSH ELISA)^26,27^, to detect RBD and full-length spike (S) reactive antibodies. Participants from Phase 1 who agreed to return for follow up serology testing (Abbott and MSH ELISA) and completion of a follow-up online survey were part of Phase 2 of the study.

### Antibody assays

The Abbott Architect assay uses a qualitative chemiluminescent microparticle immunoassay technology targeting the N antigen of the virus with a reported sensitivity of 100% (CI 95.8– 100%) and specificity of 99.6% (CI 99–99.9%)^25^. The MSH ELISA consists of an initial ELISA using serum or plasma to detect specific IgG against the RBD of SARS-CoV-2 at a single dilution, followed by quantitative titrations of presumptive positives in a confirmatory ELISA against full length SARS-CoV-2 spike protein (S)^28^. The positive result from the spike ELISA is reported as antibody at a titre of 1:80 or higher. Test performance assessment revealed that PCR+ samples were 94 % positive and all negative samples returned a negative result for 100% negative agreement^29^.

### Survey

The online survey was accessed by a unique identification number assigned to each participant, blinded to the research team to ensure confidentiality. The survey requested information on age, race/ethnicity, comorbidities, domestic/international travel and healthcare and community exposure details during and prior to both phases. The first phase collected information about symptoms of COVID-19 including their timing and duration in the preceding weeks of the blood draw^24^. The Phase 2 survey requested information on new comorbidities, persistent COVID-19 Nsymptoms (cough, shortness of breath, anosmia, ageusia, myalgia, nausea, and/or diarrhea), interim testing via antibody and/or reverse transcription polymerase chain reaction (RT-PCR) (if present) and their result (positive/negative), presence of positive SARS-CoV-2 PCR results in the preceding months, interim domestic/international travel and continued use of personal protective equipment (PPE). The risk of exposure in the healthcare setting and community exposure was determined based on CDC guidelines^30^.

### Statistical analysis

Descriptive statistics were used to summarize the baseline characteristics of the cohort and key study outcome variables. Categorical variables were compared by the Chi–squared test, while continuous variables were compared by a Student’s t-test. The spike antibody titres were described as geometric means. Correlations were calculated using standard Pearson and Spearmen correlation. Multiple linear regression was applied to determine the predictors of log10 rate of decay from Phase 1 to Phase 2 of anti-spike antibodies. A p-value of <0.05 was considered significant. All statistical analyses were performed using SPSS version 27 (IBM, USA).

## Results

For Phase 1 of our study, 500 healthcare workers underwent both PCR and serology testing. Of these, 137 were positive by for anti-N antibody (Abbott) and 142 were positive by the MSH ELISA. For the second phase 178 participants from the initial cohort consented and underwent evaluation with PCR, antibody assays (Abbott and MSH ELISA) and completed the online follow up survey. The details of patient enrolment are described in **Figure 1**. While 46 of the 178 tested subjects remained positive for the anti-N antibody (Abbott), 70 were positive by the MSH ELISA in the second phase. Anti-spike titres of the 5 subjects in the first phase were close to the cut off for positivity. Twenty-two subjects who were negative for anti-N antibody in Phase 2 had positive titres of anti-RBD and anti-spike antibodies, though lower than their Phase 1 levels. Among the subjects who participated in the Phase 1 and Phase 2 study, 68 were positive in both phases by the MSH ELISA, 110 were negative in both phases and 2 were positive only in Phase 2 with previously negative results in Phase 1.

**Figure 1.**
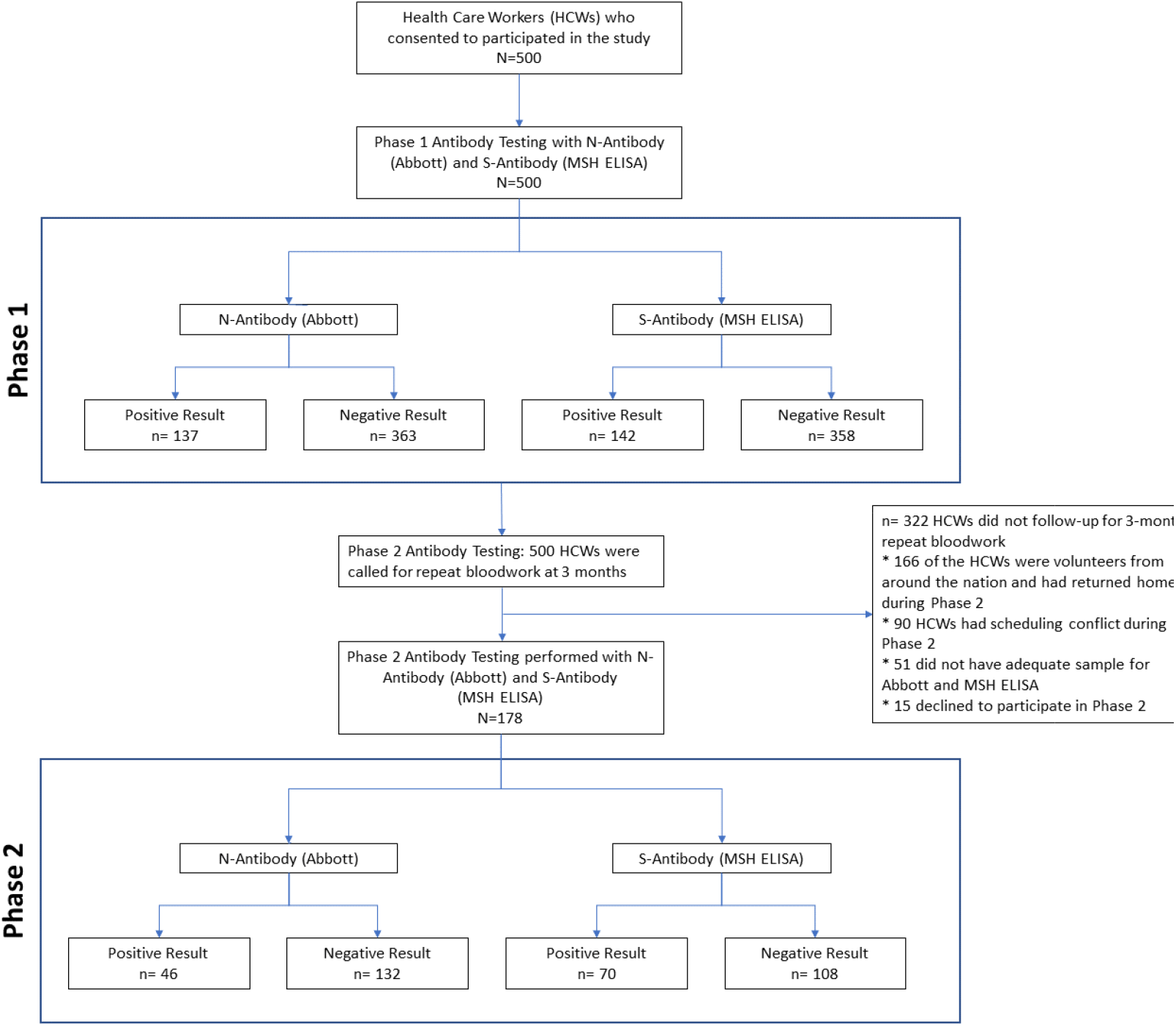
Flow Chart of patient enrollment, follow up and analysis.

The baseline characteristics of study participants who were positive by MSH ELISA in both phases (n=68) and those who were negative in both phases (n=108) are shown in **Table 1**. The mean age of the participants was 44.7±12.4 years, and 63.1% were female. Overall, 30.7% of the HCWs were Latinx, 29.5% were Asian, 16.5% were Black and 17.6% were White. COVID-19 related symptoms were present in 83.8% (57) of the subjects who were positive in both phases, while only 42.6% (46) of the subgroup who had negative antibodies in both phases admitted to symptoms prior to Phase 1. The duration of symptoms prior to Phase 1 was longer among the symptomatic positive group (48.3% for >14 days) in comparison to symptomatic negative group (17.8% for >14 days). The mean duration of symptoms to Phase 1 testing in the symptomatic positive sub cohort was 47.9 ±16.0 days. Persisting symptoms of COVID-19 were reported in 19 (27.9%) subjects from the cohort with positive antibodies in both phases.

**Table 1:**
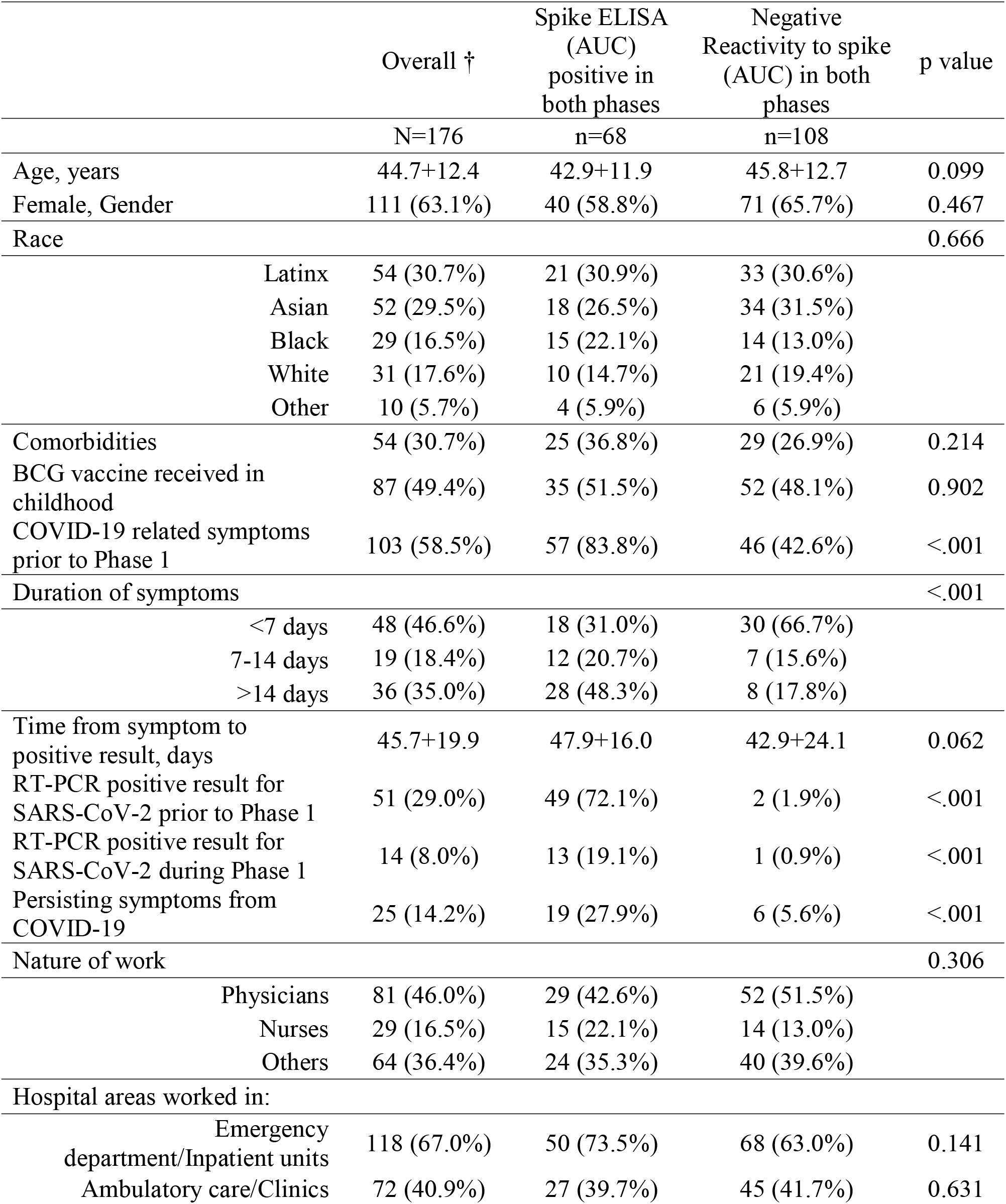
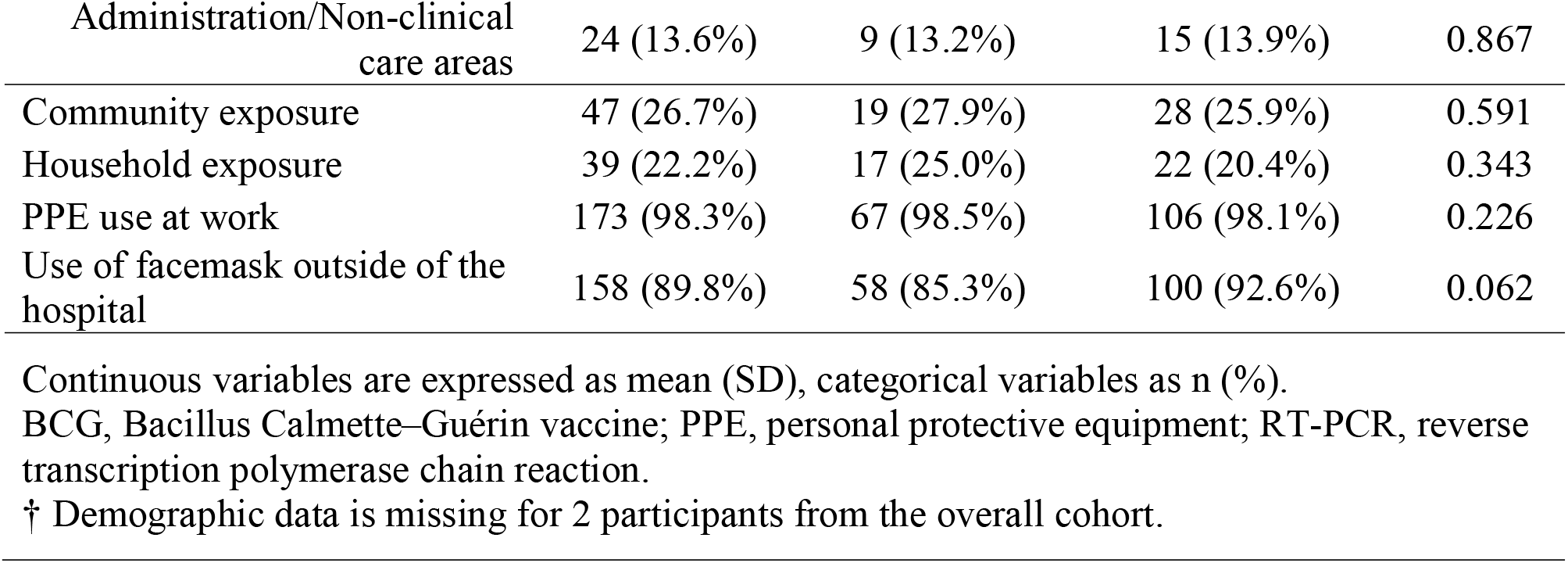
Broad characteristics among health care workers assessed for antibody reactivity to spike SARS-CoV-2 protein in Phase 1 and Phase 2

|  |  | Overall † | Spike ELISA<br>(AUC)<br>positive in<br>both phases | Negative<br>Reactivity to spike<br>(AUC) in both<br>phases | p value |
| --- | --- | --- | --- | --- | --- |
|  |  | N=176 | n=68 | n=108 |  |
| Age, years |  | 44.7+12.4 | 42.9+11.9 | 45.8+12.7 | 0.099 |
| Female, Gender |  | 111 (63.1%) | 40 (58.8%) | 71 (65.7%) | 0.467 |
| Race |  |  |  |  | 0.666 |
|  | Latinx | 54 (30.7%) | 21 (30.9%) | 33 (30.6%) |  |
|  | Asian | 52 (29.5%) | 18 (26.5%) | 34 (31.5%) |  |
|  | Black | 29 (16.5%) | 15 (22.1%) | 14 (13.0%) |  |
|  | White | 31 (17.6%) | 10 (14.7%) | 21 (19.4%) |  |
|  | Other | 10 (5.7%) | 4 (5.9%) | 6 (5.9%) |  |
| Comorbidities |  | 54 (30.7%) | 25 (36.8%) | 29 (26.9%) | 0.214 |
| BCG vaccine received in<br>childhood |  | 87 (49.4%) | 35 (51.5%) | 52 (48.1%) | 0.902 |
| COVID-19 related symptoms<br>prior to Phase 1 |  | 103 (58.5%) | 57 (83.8%) | 46 (42.6%) | <.001 |
| Duration of symptoms |  |  |  |  | <.001 |
|  | <7 days | 48 (46.6%) | 18 (31.0%) | 30 (66.7%) |  |
|  | 7-14 days | 19 (18.4%) | 12 (20.7%) | 7 (15.6%) |  |
|  | >14 days | 36 (35.0%) | 28 (48.3%) | 8 (17.8%) |  |
| Time from symptom to<br>positive result, days |  | 45.7+19.9 | 47.9+16.0 | 42.9+24.1 | 0.062 |
| RT-PCR positive result for<br>SARS-CoV-2 prior to Phase 1 |  | 51 (29.0%) | 49 (72.1%) | 2 (1.9%) | <.001 |
| RT-PCR positive result for<br>SARS-CoV-2 during Phase 1 |  | 14 (8.0%) | 13 (19.1%) | 1 (0.9%) | <.001 |
| Persisting symptoms from<br>COVID-19 |  | 25 (14.2%) | 19 (27.9%) | 6 (5.6%) | <.001 |
| Nature of work |  |  |  |  | 0.306 |
|  | Physicians | 81 (46.0%) | 29 (42.6%) | 52 (51.5%) |  |
|  | Nurses | 29 (16.5%) | 15 (22.1%) | 14 (13.0%) |  |
|  | Others | 64 (36.4%) | 24 (35.3%) | 40 (39.6%) |  |
| Hospital areas worked in: |  |  |  |  |  |
|  | Emergency<br>department/Inpatient units | 118 (67.0%) | 50 (73.5%) | 68 (63.0%) | 0.141 |
|  | Ambulatory care/Clinics | 72 (40.9%) | 27 (39.7%) | 45 (41.7%) | 0.631 |

|  |  |  |  |  |
| --- | --- | --- | --- | --- |
| Administration/Non-clinical<br>care areas | 24 (13.6%) | 9 (13.2%) | 15 (13.9%) | 0.867 |
| Community exposure | 47 (26.7%) | 19 (27.9%) | 28 (25.9%) | 0.591 |
| Household exposure | 39 (22.2%) | 17 (25.0%) | 22 (20.4%) | 0.343 |
| PPE use at work | 173 (98.3%) | 67 (98.5%) | 106 (98.1%) | 0.226 |
| Use of facemask outside of the<br>hospital | 158 (89.8%) | 58 (85.3%) | 100 (92.6%) | 0.062 |

Continuous variables are expressed as mean (SD), categorical variables as n (%).
BCG, Bacillus Calmette–Guérin vaccine; PPE, personal protective equipment; RT-PCR, reverse transcription polymerase chain reaction.
† Demographic data is missing for 2 participants from the overall cohort.

### Clinical characteristics and seropositivity to spike protein in both phases

**Table 2** describes the characteristics of the symptomatic and asymptomatic subjects who were positive for anti-spike antibody in both phases. Baseline characteristics were comparable between the groups and no difference either in the healthcare or community exposure or in the location of work (ED/Inpatient/intensive care unit, OR etc.) between the two groups was observed. Titres of anti-spike antibodies (geometric mean area under the curve (AUC)) were higher in symptomatic subjects than in asymptomatic positive subjects (6754 AUC vs. 5803 AUC) in Phase 1. However, in the Phase 2 analysis we observed marginally higher titres in the asymptomatic subgroup compared to the symptomatic subgroup (2383 AUC vs. 2198 AUC). The rate of decay was higher in the symptomatic subgroup (geometric mean 32.96 per day) compared to the asymptomatic (geometric mean 23.42 per day) suggesting delayed antibody/kinetics in the asymptomatic cohort.

**Table 2:**
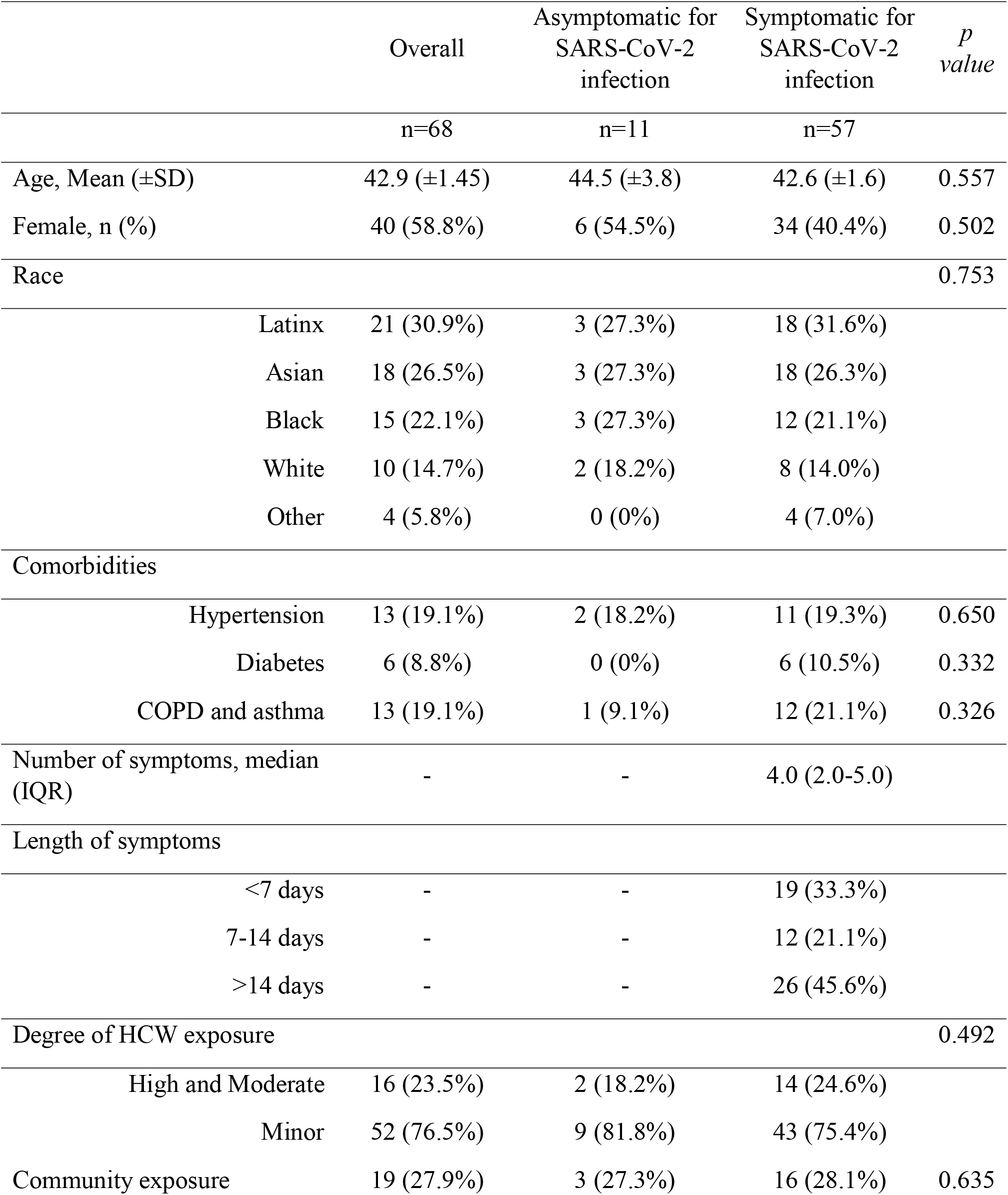
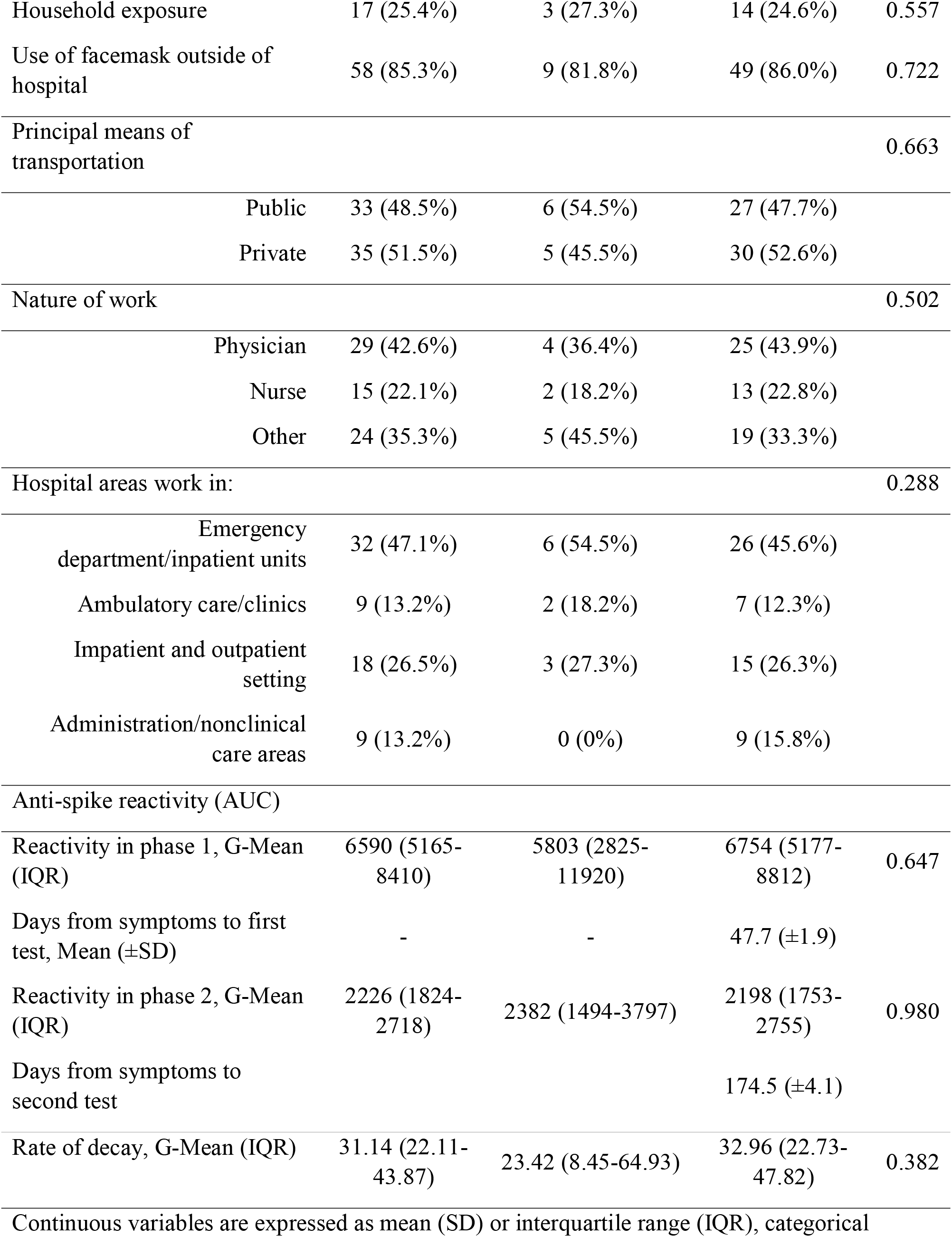
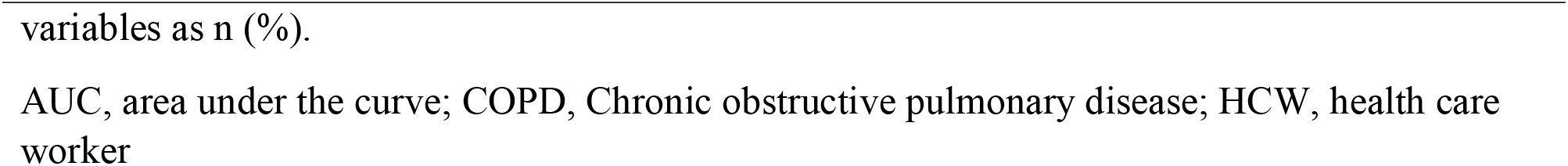
Broad characteristics among health care workers with positive antibody reactivity to SARS-CoV-2 spike in both phases

|  | Overall | Asymptomatic for<br>SARS-CoV-2<br>infection | Symptomatic for<br>SARS-CoV-2<br>infection | *p*<br>*value* |
| --- | --- | --- | --- | --- |
|  | n=68 | n=11 | n=57 |  |
| Age, Mean (±SD) | 42.9 (±1.45) | 44.5 (±3.8) | 42.6 (±1.6) | 0.557 |
| Female, n (%) | 40 (58.8%) | 6 (54.5%) | 34 (40.4%) | 0.502 |
| Race |  |  |  | 0.753 |
| Latinx | 21 (30.9%) | 3 (27.3%) | 18 (31.6%) |  |
| Asian | 18 (26.5%) | 3 (27.3%) | 18 (26.3%) |  |
| Black | 15 (22.1%) | 3 (27.3%) | 12 (21.1%) |  |
| White | 10 (14.7%) | 2 (18.2%) | 8 (14.0%) |  |
| Other | 4 (5.8%) | 0 (0%) | 4 (7.0%) |  |
| Comorbidities |  |  |  |  |
| Hypertension | 13 (19.1%) | 2 (18.2%) | 11 (19.3%) | 0.650 |
| Diabetes | 6 (8.8%) | 0 (0%) | 6 (10.5%) | 0.332 |
| COPD and asthma | 13 (19.1%) | 1 (9.1%) | 12 (21.1%) | 0.326 |
| Number of symptoms, median (IQR) | - | - | 4.0 (2.0-5.0) |  |
| Length of symptoms |  |  |  |  |
| <7 days | - | - | 19 (33.3%) |  |
| 7-14 days | - | - | 12 (21.1%) |  |
| >14 days | - | - | 26 (45.6%) |  |
| Degree of HCW exposure |  |  |  | 0.492 |
| High and Moderate | 16 (23.5%) | 2 (18.2%) | 14 (24.6%) |  |
| Minor | 52 (76.5%) | 9 (81.8%) | 43 (75.4%) |  |
| Community exposure | 19 (27.9%) | 3 (27.3%) | 16 (28.1%) | 0.635 |
| Household exposure | 17 (25.4%) | 3 (27.3%) | 14 (24.6%) | 0.557 |
| Use of facemask outside of hospital | 58 (85.3%) | 9 (81.8%) | 49 (86.0%) | 0.722 |
| Principal means of transportation |  |  |  | 0.663 |
| Public | 33 (48.5%) | 6 (54.5%) | 27 (47.7%) |  |
| Private | 35 (51.5%) | 5 (45.5%) | 30 (52.6%) |  |
| Nature of work |  |  |  | 0.502 |
| Physician | 29 (42.6%) | 4 (36.4%) | 25 (43.9%) |  |
| Nurse | 15 (22.1%) | 2 (18.2%) | 13 (22.8%) |  |
| Other | 24 (35.3%) | 5 (45.5%) | 19 (33.3%) |  |
| Hospital areas work in: |  |  |  | 0.288 |
| Emergency department/inpatient units | 32 (47.1%) | 6 (54.5%) | 26 (45.6%) |  |
| Ambulatory care/clinics | 9 (13.2%) | 2 (18.2%) | 7 (12.3%) |  |
| Inpatient and outpatient setting | 18 (26.5%) | 3 (27.3%) | 15 (26.3%) |  |
| Administration/nonclinical care areas | 9 (13.2%) | 0 (0%) | 9 (15.8%) |  |
| Anti-spike reactivity (AUC) |  |  |  |  |
| Reactivity in phase 1, G-Mean (IQR) | 6590 (5165-8410) | 5803 (2825-11920) | 6754 (5177-8812) | 0.647 |
| Days from symptoms to first test, Mean (±SD) | - | - | 47.7 (±1.9) | |
| Reactivity in phase 2, G-Mean (IQR) | 2226 (1824-2718) | 2382 (1494-3797) | 2198 (1753-2755) | 0.980 |
| Days from symptoms to second test | | | 174.5 (±4.1) | |
| Rate of decay, G-Mean (IQR) | 31.14 (22.11-43.87) | 23.42 (8.45-64.93) | 32.96 (22.73-47.82) | 0.382 |
| Continuous variables are expressed as mean (SD) or interquartile range (IQR), categorical |  |  |  |  |

variables as n (%).
AUC, area under the curve; COPD, Chronic obstructive pulmonary disease; HCW, health care worker

### Phase 1 anti-spike antibody titre and clinical correlations

A Pearson’s product and Spearman’s rank-order correlation was run to assess the relationship between cohort characteristics including age, gender, comorbidities, number of symptoms of COVID-19, healthcare exposure and Phase 1 anti-spike titres in our cohort (**Figure 2**). One hundred-forty-three subjects with a positive test in Phase 1 were included in the analysis. Scatter plot analysis showed a monotonic relationship between the variables. A statistically significant weak positive correlation was observed between age and Phase 1 anti-spike antibody titres (R=0.269, p<0.005). Moderate positive correlation was present between presence of fever (R=0.319, p<0.005), number of symptoms (R=0.310, p<0.005) and days of symptoms (R=0.434, p<0.005) and anti-spike antibody titre; and weak positive correlation was observed with upper respiratory symptoms (R=0.278, p<0.005) and gastrointestinal (GI) symptoms (R=0.204, p<0.05) with anti-spike antibody titres.

**Figure 2.**
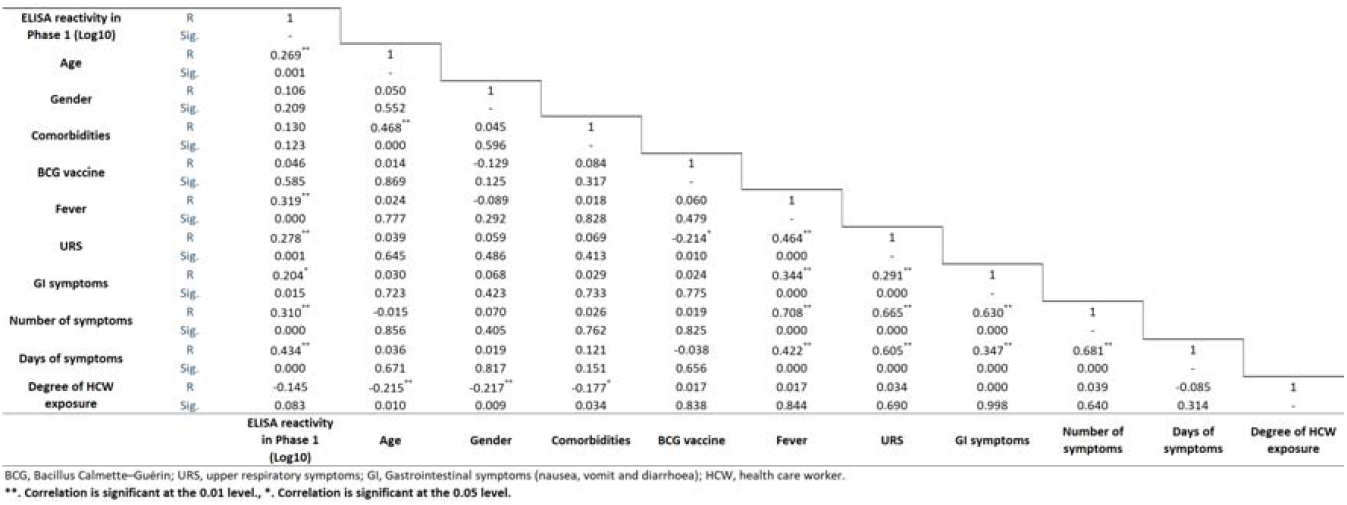
Simple correlation analysis of HCW with positive reactivity for anti-spike antibody in Phase 1 with baseline characteristics and symptoms.

### Correlation of rate of decay of anti-spike antibody titres from Phase 1 to Phase 2 and clinical characteristics

Results of Pearson correlation to assess the relationship between cohort characteristics including Phase 1 anti-spike antibody titres, age, gender, comorbidities, symptoms of COVID-19, number of symptoms of COVID-19, healthcare exposure and decay of anti-spike titres between the two phases in our cohort is shown in **Figure 3**. A strong positive statistically significant correlation was observed between Phase 1 titers and decay of anti-spike antibody titres between the two phases (R=0.898, p<0.000). Medium positive correlation was observed between presence of fever (R=0.428, p<0.001), GI symptoms (R=0.340, p<0.011), number of symptoms (R=0.357, p<0.007), duration of symptoms (R=0.469, p<0.000) with decay of anti-spike antibody titres between the two phases respectively.

**Figure 3.**
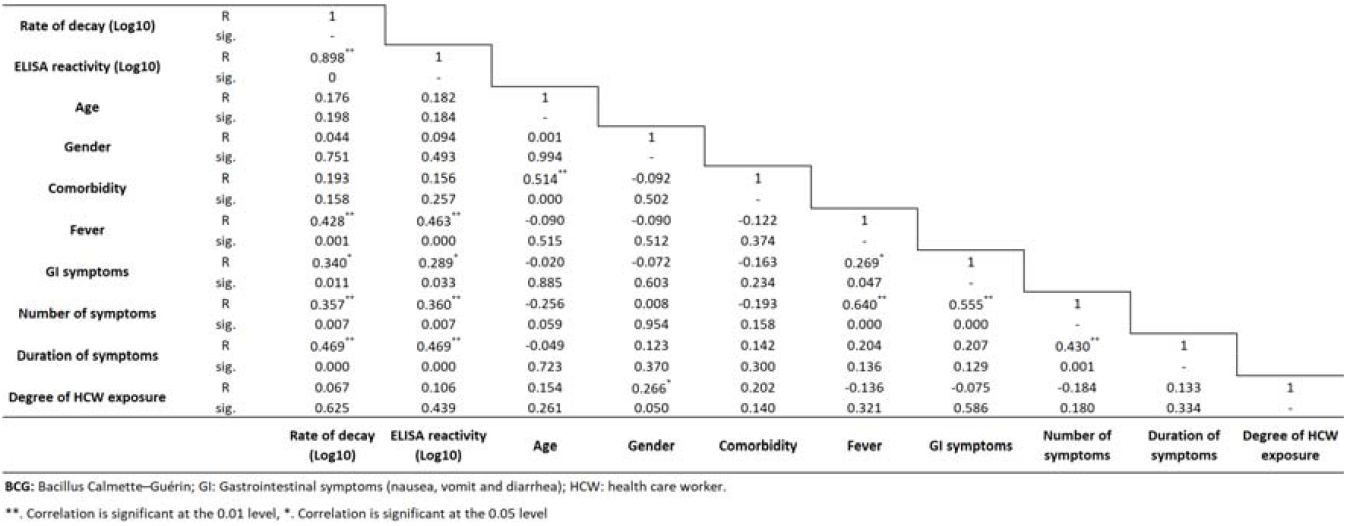
Simple correlation analysis of rate of decay of anti-spike antibodies between both phases with baseline characteristics and symptoms.

A pairwise comparison was performed between rate of decay of anti-spike antibody titres and patient characteristics (**Figure 4**). Rate of decay by gender was comparable (male; 30.73 AUC/day vs. female;34.68 AUC/day, p=0.413). Asian (86.0 AUC/day) race showed higher rate of decay compared with White (7.2 AUC/day) and Black (19.61 AUC/day) individuals; while Latinx (47.28 AUC/day) race had higher rate of decay compared with White (7.2 AUC/day) individuals. Subjects with fever had a higher rate of decay than those who did not report fever (53.08 AUC/day vs.16.14 AUC/day, p=0.002). Similarly subjects with GI symptoms had a higher rate of decay than those without (55.81 AUC/day vs.21.94 AUC/day, p=0.019). Subjects with symptoms restricted to less than seven days demonstrated a lower decay rate compared with symptomatic subjects over 7-14 days (13.60 AUC/day vs. 36.12 AUC/day, p=0.046) and when compared with symptomatic subjects with more than 14 days (13.60 AUC/day vs. 59.72 AUC/day, p=0.001). This finding was statistically significant. No difference was found when degree of exposure (High/Moderate: 28.18 AUC/day vs. Mild: 34.78 AUC/day, p=0.395) or job role (physician: 29.57 AUC/day vs. nurse: 53.59 AUC/day vs. Other: 26.83 AUC/day; p=0.361) was compared to rate of decay.

**Figure 4.**
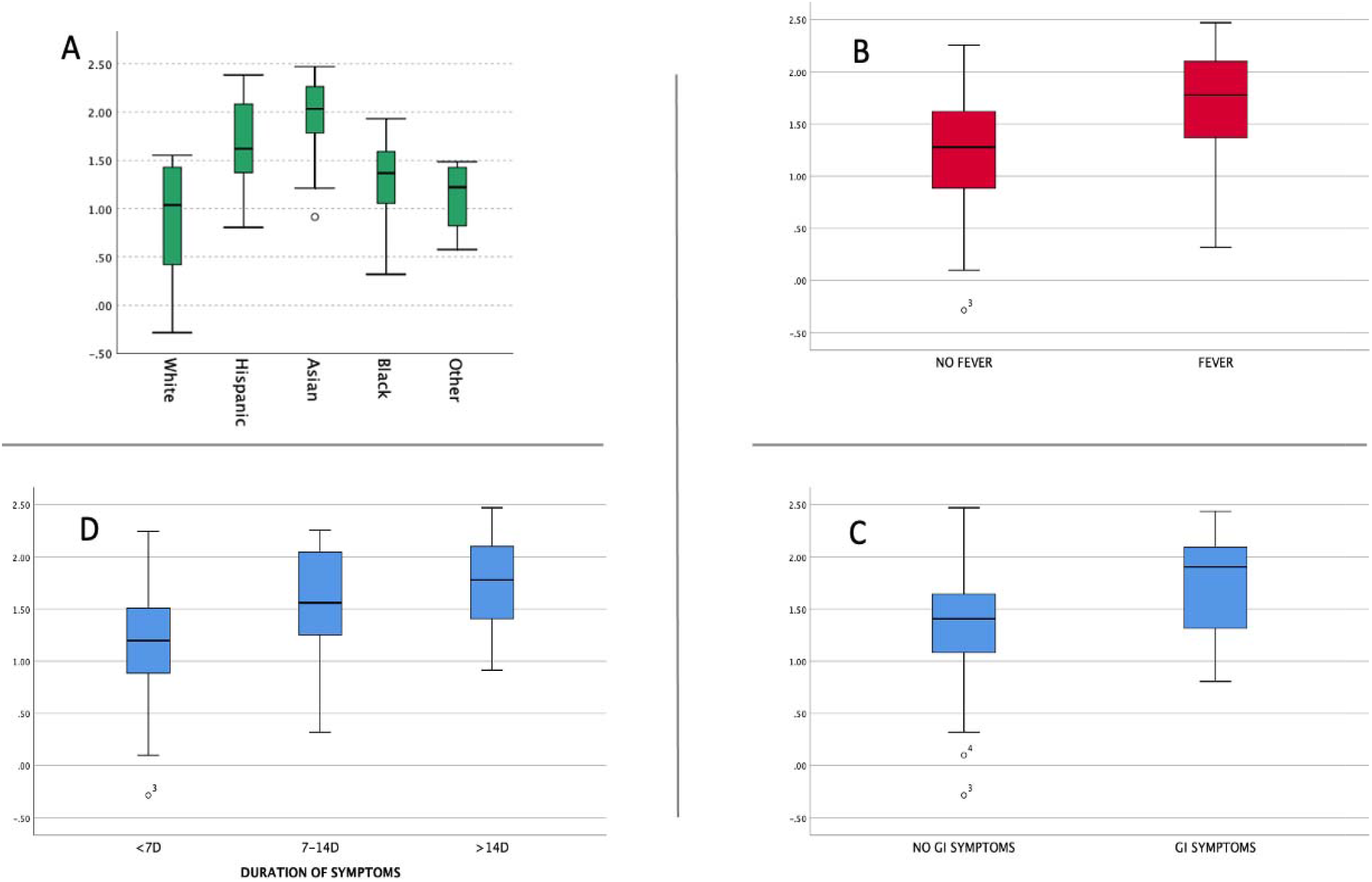
Paired comparison between rate of decay of anti-spike antibody titres and patient characteristics.

### Predictors of rate of decay from Phase 1 to Phase 2 of anti-spike antibodies

Multiple linear regression analysis to predict the rate of decay with respect to age, Bacillus Calmette Guerin vaccination, number of symptoms, and Phase 1 (log10) anti-spike antibody titres is shown in **Table 3**. On the basis of a linear regression model that included the participants age, history of BCG vaccination, total number of COVID-19 symptoms and the Phase 1 concentration of log 10 spike antibody titres, the estimated change (decay) was 23.6 AUC/day when age was centred at median (42.6 years), there was positive history of BCG vaccination, the total number of COVID-19 symptoms were centred at a median of 4, and the geometric mean of the log_10_ spike antibody titre was 3.78.

**Table 3:** Multiple linear regression analysis of rate of decay for anti-spike antibodies between Phase 1 and Phase 2

| Rate of decay<br>(log10) | *B* | 95.0% CI for *B* | | *SE B* | $\beta$ | $R^2$ | $\Delta R^2$ |
| --- | --- | --- | --- | --- | --- | --- | --- |
|  |  | LL | UL |  |  |  |  |
| **Model** |  |  |  |  |  | 0.83 | 0.82 |
| **Constant** | -3.203** | -3.647 | -2.759 | .222 |  |  |  |
| **Age (per 10-year change)** | .014 | -.005 | .007 | .002 | .030 |  |  |
| **BCG vaccination** | .131** | .030 | .310 | .046 | .121 |  |  |
| **Number of symptoms** | .013 | -.029 | .060 | .012 | .050 |  |  |
| **ELISA reactivity (Log10)** | 1.159** | 1.050 | 1.419 | .059 | .916 |  |  |

*B*: Unstandardized regression coefficient; CI: confidence interval; LL: lower limit; UL: upper limit; *SE B*: standard error of the coefficient; $\beta$ : standardized coefficient; $R^2$ : coefficient of determination; $\Delta R^2$ : adjusted $R^2$ .
\*\* $P < 0.05$
BCG, Bacillus Calmette–Guérin vaccine

## Discussion

With the COVID-19 pandemic showing no signs of abating, healthcare workers at the epicentre are at risk of infection due to occupational exposure as well as community exposure. Sero-surveillance is the foundation for determining the scale and rate of exposures. With a multitude of serological assays getting emergency use approval from FDA, interpretation of the results of these assays and their clinical significance remains challenging. It is critical to understand the timing of the antibody response for acute interpretation. Confidence in analytical specificity of the assay is a critical requirement in measurement of the specific antibody responses. Recent studies have confirmed that anti spike titres especially anti-RBD titres can serve as surrogates for virus neutralization^31,32^. The Abbott SARS-CoV-2 IgG assay that targets antibodies to the nucleoprotein has a reported specificity and sensitivity of greater than 99% at 14 days or more following symptom onset and these measurements are not indicative or correlated to virus neutralization titres^33^. In comparison, the MSH ELISA targets the full-length S protein including RBD, a major target for neutralizing antibodies and has demonstrated excellent correlation to virus neutralization^11,26^. Longitudinal measurements of antibody levels have revealed that anti-N and anti S IgG antibodies continue to increase until the third week post symptoms and an approach that combines the detection of both of these antibodies would precisely detect almost 100% of all infectious exposures^34^. In our study, the mean number of days after symptoms to testing in Phase 1 was 47 days suggesting a higher likelihood of accuracy of the utilized assay.

Longitudinal blood sampling among HCWs working at a public hospital in the epicentre of the pandemic in NYC allowed for analysis of kinetics of anti-S and anti-N antibody responses. At two months after the first surge of infections, anti-N antibodies were detected in 27% and anti-S antibodies in 28% of participating HCWs. After an interval of four months, it is not surprising to note that among the participants who returned, 26% remained positive for anti N antibodies, while 31% of the previously anti-N antibody positive subjects tested negative in phase 2. On the other hand, a similar analysis of the anti-S antibodies levels, confirmed that all the previously positive retested subjects continued to remain positive, albeit with lower titres.

COVID-19 related symptoms were significantly associated with positive anti-spike antibodies in both phases, with a similar association with longer duration (>14 days) of symptoms. Previous studies have demonstrated a lower level of IgG response among patients without symptoms or with mild symptoms compared to those with severe and critical disease^35,36^. A comparison of symptomatic versus asymptomatic subjects who tested positive for anti-spike antibodies in both phases, confirmed that the rate of decay of anti-spike antibody titres were faster in the symptomatic cohort than the asymptomatic subjects, which was seen also in the anti-N antibody kinetics. However, we observed a faster decay in this group with a lower titre of anti-spike antibodies in Phase 2 compared to the asymptomatic cohort (though the difference was not statistically significant). This could additionally be supported by the finding of fever and GI symptoms contributing to faster decay. Similar results of decreasing neutralizing antibody titre in symptomatic than asymptomatic patients were observed by Choe *et al*.^37^

Positive correlations for age, presence of fever, upper respiratory symptoms, GI symptoms, total number and duration of symptoms was observed with increased levels of anti-spike titres at Phase 1. Similar results of neutralizing antibody titres were also observed by Boonyaratanakornkit *et al*. wherein they showed higher levels of neutralizing antibody titres were significantly associated with male gender, older adults, higher disease severity and shorter interval from recovery^38^. Based on a linear regression model with age centred at median (42.6 years), positive history of BCG vaccination, the total number of COVID-19 symptoms centred at a median of 4, and the geometric mean of the log10 anti-spike antibody titre at 3.78, we observed that the rate of decay of these antibody titres was 23.6 AUC/day. Evaluation of other characteristics with rate of decay between Phase 1 and Phase 2 showed a faster reduction in titres in Asian participants and in those with fever and GI symptoms. A slower decrease was noted among patients with shorter duration (<7 days) of symptoms, with no other significant correlation noted with any other baseline demographics or clinical characteristics.

As described above, higher antibody titers are associated with a larger number of symptoms, longer duration of symptoms and – as described by others as well – disease severity in general. We also found that higher initial antibody titers were associated with faster antibody decay during the two time points. Initial antibody responses are driven by short lived plasmablasts, which decay after a few days after producing massive amounts of antibody. IgG has a relatively long half-life of approximately three weeks, but decay is inevitable since the plasmablasts initially producing it disappear. Usually, titers then drop until they reach relatively stable levels of antibody which are maintained by long-lived plasma cells in the bone marrow^19^. The two time points described in this study represent the initial peak response and likely the stable level after the initial decay. We found that individuals with higher initial titers had a faster decay rate during the observation period meaning the difference between peak and stable, long-lived antibody levels were larger. This indicates that there is likely no direct correlation between the magnitude of the initial expansion of plasmablasts and the number of long-lived plasma cells that migrate to the bone marrow.

Our study has the following limitations: First being a single center study with a small convenience sampling that included a smaller number of participants in Phase 2 of the study. Following the pandemic, the HCWs who volunteered from around the country were transferred back and lost to follow-up, which did decrease the overall sample size, but the rates of positive and negative results remained proportional. Second, the likelihood of a recall bias in the participant’s responses on the online survey may exist. Lastly, as a cross-sectional seroprevalence study the findings can underestimate rates of prior infections based on timing of the testing given that antibodies are only transiently detectable following infection. In conclusion, findings from this study are similar to other studies that have reported that higher magnitude of anti-spike titres may correlate with protection against reinfection, in spite of the observed decay in the antibody levels^20,21^. Nevertheless, further studies to evaluate the longevity of immunity, especially in context of widespread administration of spike-based vaccine among HCWs would be important in predicting herd immunity to COVID-19 infections.

## Data Availability

All the data that had been collected has been presented in the manuscript.

## Acknowledgements

The authorship is structured in first-last-author-emphasis. We thank the COVID-19 Testing Tent staff for their invaluable assistance with this study with testing and accommodation of study participants for Phase 2: Patrick C McNeil PA-C MPAS; Aney D Patel PA-C MPAS; and Megan Corley DNP ANP-BC. We thank the Nursing staff for their commitment and support with the workflow, especially, Karen Philip and Kenisha Williamson with others; additionally, the Registration (Lexus Gonzalez and Taj Washington) and Patient Care Associate (Eva Penn) that assisted the research team. We also thank the Occupational Health Services for accommodating the research team during project completion. The Clinical Laboratory staff played a part in allocating resources for Abbott Architect usage: Dior Ndao and Ayman Elshamshery. We thank the study participants, who were essential health care workers, who volunteered to follow-up in this protocol.

## Contributions

V.M., M.A.S. and U.V. designed the study. J.M.C., M.A.S., B.Y., V.P.G. analysed the data. E.V. and M.G. assisted with participant follow-up and coordination with the assistance of A.P., U.V., V.M., and M.A.S. The Mount Sinai Health System team, J.M.C. and F.K., performed the measurements for anti-Spike and Anti-RBD antibodies. V.M., U.V. and A.P. were responsible for the clinical care of the research participants and supervised the day-to-day operation and coordination of the study by M.K., V.D., M.A.S., B.Y., V.P.G., M.G., E.V., and M.G. V.M. and F.K wrote the manuscript and is the guarantor of this work and has full access to all data in the study and takes responsibility for the integrity of the data and the accuracy of the data analysis, with the assistance of J.C.Q., M.A.S., B.Y., V.P.G., and M.G. All authors critically revised the draft and approved the final manuscript.

## References

1. Ou X, Liu Y, Lei X, et al. Characterization of spike glycoprotein of SARS-CoV-2 on virus entry and its immune cross-reactivity with SARS-CoV. Nat Commun. 2020;11(1):1620.

2. Walls AC, Park YJ, Tortorici MA, Wall A, McGuire AT, Veesler D. Structure, Function, and Antigenicity of the SARS-CoV-2 Spike Glycoprotein. Cell. 2020;181(2):281–292.

3. Premkumar L, Segovia-Chumbez B, Jadi R, et al. The receptor binding domain of the viral spike protein is an immunodominant and highly specific target of antibodies in SARS-CoV-2 patients. Sci Immunol. 2020;5(48):eabc8413.

4. Ni L, Ye F, Cheng ML, et al. Detection of SARS-CoV-2-Specific Humoral and Cellular Immunity in COVID-19 Convalescent Individuals. Immunity. 2020;52(6):971–977

5. Iyer AS, Jones FK, Nodoushani A, et al. Persistence and decay of human antibody responses to the receptor binding domain of SARS-CoV-2 spike protein in COVID-19 patients. Sci Immunol. 2020; 5(52):eabe0367.

6. Cong Y, Ulasli M, Schepers H, et al. Nucleocapsid Protein Recruitment to Replication-Transcription Complexes Plays a Crucial Role in Coronaviral Life Cycle. J Virol. 2020;94(4):e01925–19.

7. Burbelo PD, Riedo FX, Morishima C, et al. Detection of Nucleocapsid Antibody to SARS-CoV-2 is More Sensitive than Antibody to Spike Protein in COVID-19 Patients. medRxiv [Preprint]. 2020;2020.04.20.20071423

8. Long QX, Liu BZ, Deng HJ, Wu GC, Deng K, Chen YK. Antibody responses to SARS-CoV-2 in patients with COVID-19. Nat Med. 2020;26(6):845–848.

9. İnandiklioğlu N, Akkoc T. Immune Responses to SARS-CoV, MERS-CoV and SARS-CoV-2. Adv Exp Med Biol. 2020;1288:5–12.

10. Chaudhuri S, Thiruvengadam R, Chattopadhyay S, et al Comparative evaluation of SARS-CoV-2 IgG assays in India. J Clin Virol. 2020;131:104609.

11. Wajnberg A, Amanat F, Firpo A, et al. Robust neutralizing antibodies to SARS-CoV-2 infection persist for months. Science. 2020;370(6521):1227–1230.

12. Crawford KHD, Dingens AS, Eguia R, et al. Dynamics of neutralizing antibody titers in the months after SARS-CoV-2 infection. J Infect Dis. 2020:jiaa618.

13. Luchsinger LL, Ransegnola BP, Jin DK, et al. Serological Assays Estimate Highly Variable SARS-CoV-2 Neutralizing Antibody Activity in Recovered COVID-19 Patients. J Clin Microbiol. 2020;58(12)

14. Iyer AS, Jones FK, Nodoushani A, et al. Dynamics and significance of the antibody response to SARS-CoV-2 infection. medRxiv. 2020;2020.07.18.20155374.

15. Isho B, Abe KT, Zuo M, et al. Persistence of serum and saliva antibody responses to SARS-CoV-2 spike antigens in COVID-19 patients. Sci Immunol. 2020;5(52):p eabe5511.

16. Wu F, Wang A, Liu m, et al. Neutralizing antibody responses to SARS-CoV-2 in a COVID-19 recovered patient cohort and their implications. medRxiv, 2020;2020.03.30.20047365.

17. Seow J, Graham C, Merrick B, et al. Longitudinal observation and decline of neutralizing antibody responses in the three months following SARS-CoV-2 infection in humans. Nat Microbiol. 2020;5(12):1598–1607.

18. Grandjean L, Saso A, Torres A, et al. Humoral Response Dynamics Following Infection with SARS-CoV-2. medRxiv, 2020;2020.07.16.20155663.

19. Ellebedy A, Turner J, Kim W, et al. SARS-CoV-2 infection induces long-lived bone marrow plasma cells in humans. Res Sq [Preprint]. 2020:rs.3.rs-132821.

20. Lumley SF, O’Donnell D, Stoesser NE, et al. Antibody Status and Incidence of SARS-CoV-2 Infection in Health Care Workers. N Engl J Med. 2021;384(6):533–540.

21. Hall V, Foulkes S, Charlett A, et al. Do antibody positive healthcare workers have lower SARS-CoV-2 infection rates than antibody negative healthcare workers? Large multi-centre prospective cohort study (the SIREN study), England: June to November 2020. medRxiv, 2021;2021.01.13.21249642.

22. Self WH, Tenforde MW, Stubblefield WB, et al. Seroprevalence of SARS-CoV-2 Among Frontline Health Care Personnel in a Multistate Hospital Network. 13 Academic Medical Centers, April-June 2020. MMWR Morb Mortal Wkly Rep. 2020;69(35):1221–1226.

23. Moscola J, Sembajwe G, Jarrett M, et al. Prevalence of SARS-CoV-2 Antibodies in Health Care Personnel in the New York City Area. JAMA. 2020;324(9):893–895.

24. Venugopal U, Jilani N, Rabah S, et al. SARS-CoV-2 seroprevalence among health care workers in a New York City hospital: A cross-sectional analysis during the COVID-19 pandemic. Int J Infect Dis. 2021;102:63–69

25. H Hamilton F Muir P, Attwood M, et al. Kinetics and performance of the Abbott architect SARS-CoV-2 IgG antibody assay. J Infect. 2020;81(6):e7–e9.

26. Amanat F, Stadlbauer D, Strohmeier S, et al. A serological assay to detect SARS-CoV-2 seroconversion in humans. Nat Med. 2020;26(7):1033–1036.

27. Stadlbauer D, Amanat F, Chromikova V, et al. SARS-CoV-2 Seroconversion in Humans: A Detailed Protocol for a Serological Assay, Antigen Production, and Test Setup. Curr Protoc Microbiol. 2020;57(1):e100.

28. Amanat F, Stadlbauer D, Strohmeier S, et al. A serological assay to detect SARS-CoV-2 seroconversion in humans. Nat Med. 2020;26(7):1033–1036.

29. Stadlbauer D, Tan J, Jiang K, et al. Repeated cross-sectional sero-monitoring of SARS-CoV-2 in New York City. Nature. 2021;590(7844):146–150.

30. Garcia M, Lipskiy N, Tyson J, Watkins R, Esser ES, Kinley T. Centers for Disease Control and Prevention 2019 novel coronavirus disease (COVID-19) information management: addressing national health-care and public health needs for standardized data definitions and codified vocabulary for data exchange. J Am Med Inform Assoc. 2020;27(9):1476–1487

31. Therrien C, Serhir B, Bélanger-Collard M, et al. Multicenter Evaluation of the Clinical Performance and the Neutralizing Antibody Activity Prediction Properties of ten high throughput serological assays used in Clinical Laboratories. J Clin Microbiol. 2020;JCM.02511-20.

32. Muecksch F, Wise H, Batchelor B, et al. Longitudinal analysis of clinical serology assay performance and neutralising antibody levels in COVID19 convalescents. medRxiv, 2020;2020.08.05.20169128.

33. Rosadas C, Randell P, Khan M, McClure MO, Tedder RS. Testing for responses to the wrong SARS-CoV-2 antigen? Lancet. 2020;396(10252):e23.

34. Sun B, Feng Y, Mo X, et al. Kinetics of SARS-CoV-2 specific IgM and IgG responses in COVID-19 patients. Emerg Microbes Infect. 2020;9(1):940–948.

35. Long QX, Tang XJ, Shi QL, et al. Clinical and immunological assessment of asymptomatic SARS-CoV-2 infections. Nat Med. 2020; 26(8):1200–1204.

36. Wang Y, Zhang L, Sang L, et al. Kinetics of viral load and antibody response in relation 1. to COVID-19 severity. J Clin Invest. 2020;130(10):5235–5244.

37. Choe PG, Kang CK, Suh HJ, et al. Waning Antibody Responses in Asymptomatic and Symptomatic SARS-CoV-2 Infection. Emerg Infect Dis. 2021;27(1):327–329.

38. Boonyaratanakornkit J, Morishima C, Selke S, et al. Clinical, laboratory, and temporal predictors of neutralizing antibodies to SARS-CoV-2 after COVID-19. medRxiv. 2020; 2020.10.06.20207472.

